# Prenatal exposure to genocide accelerates epigenetic aging as measured in second-generation clocks among young adults

**DOI:** 10.1101/2024.10.01.24314372

**Authors:** Glorieuse Uwizeye, Luisa M. Rivera, Hannah G. Stolrow, Brock C. Christensen, Julienne N. Rutherford, Zaneta M. Thayer

**Affiliations:** Arthur Labatt Family School of Nursing, Western University, London, ON, Canada; Department of Anthropology, Dartmouth College, Hanover, NH, USA; Department of Epidemiology, Geisel School of Medicine, Dartmouth College, Lebanon, ON, USA; Advanced Nursing Practice and Science Division, College of Nursing, University of Arizona, Tucson, AZ, USA

**Keywords:** Epigenetic clocks, Prenatal exposure to stress, Genocide against the Tutsi, Adverse early life experience, Rape, Rwanda

## Abstract

Prenatal exposure to trauma, including genocide and maternal rape, and adverse childhood experiences (ACEs), are associated with lifespan reduction. We evaluated whether prenatal exposure to genocide or genocidal rape, and ACEs among individuals conceived during the 1994 genocide against Tutsi in Rwanda were associated with differences in age acceleration in three first-generation (Horvath, Hannum, PhenoAge) and four second-generation epigenetic aging clocks (GrimAge, DunedinPace, YingDamAge, YingAdaptAge), given the association between biological aging and mortality. No differences in age acceleration were observed with first-generation age clocks. However, age acceleration was associated with prenatal exposure to extreme stress for all second-generation clocks, with the greatest acceleration observed in the genocidal rape conception group. For YingDamAge clock, acceleration effects were strengthened after inclusion of ACEs. We suggest that prenatal trauma exposure is associated with epigenetic age acceleration. Second-generation clocks may more accurately capture these relationships.

## INTRODUCTION

Genocide has profound and long-lasting effects on humanity. In Rwanda, more than 1,000,000 lives were lost in a period of 100 days in the 1994 genocide against the Tutsi^1^. The genocide not only impacted the lives of those who directly experienced it but also those who were indirectly exposed while in utero. Our previous studies demonstrated that young adults conceived during the genocide against the Tutsi had worse mental and physical function and higher post- traumatic stress disorder (PTSD) scores, anxiety, depression, pain intensity, and sleep disturbance compared to age- and sex-matched young adults who were not prenatally exposed to the genocide^2^. Other studies conducted in Rwanda also reported significantly higher scores of PTSD, depression and lower cortisol levels among Rwandans prenatally exposed to genocide when compared with those whose parents were living outside the country during the time of genocide^3, 4^.

Rape was used as a systematic weapon during the 1994 genocide and affected approximately 350,000 women, of whom only one in six survived^2, 5^. While the exact number of children born as a result of genocidal rape will never be known, the total is estimated between 2,000-10,000 ^5,6^. Individuals conceived during the genocide, including those conceived through rape, were exposed to this trauma during the first trimester, which is a critical stage of development. For those conceived via genocidal rape, stress related to their birth origins extends beyond the acute period of genocide and continues throughout childhood^7, 8^. For example, studies conducted in Rwanda and the former Yugoslavia reported that children born of genocidal rape face physical and emotional abuse from family and community members and endure poverty and other socioeconomic hardships^7, 9^. This can manifest in a significantly higher likelihood of experiencing adverse childhood experiences ^2^.

While research has demonstrated associations between prenatal exposure to genocide and mental and physical health outcomes^2, 10^, it is possible that prenatal exposure to genocide and rape also impacted various aspects of biological regulation, including patterns of DNA methylation. We previously reported that individuals who were born of genocidal rape, relative to controls, had DNA methylation that varied at CpGs in *BDNF* and *SLC6A4*, and methylation in these sites was associated with adult mental health outcomes^11^. While suggestive, site-specific DNA methylation has not proven to be a useful prognosticator of health; by contrast, DNA methylation-based aging estimators have been associated with mental health treatment outcomes^12^, cancer prognosis, and chronic disease mortality^13^. There are different machine-learning algorithms to estimate biological aging. First-generation epigenetic age estimators were created by training neural-net models to predict age by comparing the methylation status at CpG sites present on arrays with chronological age (e.g., Horvath^14^, Hannum^15^); age acceleration was conceptualized as the positive difference (either raw or residualized) between estimated and actual age. Such estimators have been critiqued for low test-retest reliability,^16^ a lack of generalizability beyond their training data,^17^ and a limited ability to capture biological processes and/or epigenetic patterns related to healthy aging and longevity. In response, so-called “second generation” epigenetic clocks have included phenotypic measures known to associate with biological aging (e.g. PhenoAge) as well as known longitudinal mortality and longevity data (GrimAge, DunedinPACE) and most recently, causally-constrained epigenetic markers of adaptive aging or longevity (YingAdaptAge) and/or decreased lifespan or damage-related aging (YingDamAge)^18^. In this paper, we categorize PhenoAge as a first-generation clock; while it does include serum biomarkers associated with poor health, it does not include longitudinal mortality data (e.g. GrimAge), longitudinal age-related decline (DunedinPACE) or longitudinal age-related morbidity/mortality and longevity outcomes (YingDamAge, YingAdaptAge) in its training data (see Supplemental Table 1). Though effect sizes vary depending on the clock used, accelerated epigenetic age has been associated with decreased lifespan and “healthspan”^19^. Notably, early life adversity (ELA, e.g. both prenatally and in childhood) has been associated with accelerated epigenetic age, although these associations frequently vary depending on the epigenetic clock used and the way adversity is measured^20^. For example, in a systematic review of ELA and epigenetic aging in the Horvath and Hannum clocks, experiences of threat (vs. deprivation) were most related to biological aging^21^ in children. In a separate analysis among Congolese newborns, prenatal exposure to general trauma and war trauma was found to be associated with accelerated epigenetic age in the Hannum extrinsic age (but not PhenoAge or GrimAge) clocks^22^. Lower birth weight has also been associated with accelerated epigenetic aging in the Hannum, DNAmPhenoAge, DunedinPoAm, and DNAmTL (but not GrimAge) in young male, but not female, adults in the Philippines^23^. Here, we use DNA methylation array data from our previously described cohort of individuals conceived during the 1994 genocide against the Tutsi^11^ to evaluate epigenetic aging using all seven published epigenetic age estimators.

### Hypotheses

The primary aim of this study was to evaluate whether different patterns of prenatal exposure to maternal stress are associated with epigenetic age. We evaluated three groups, 1) single-exposed - maternal stress related to genocide; 2) double-exposed - maternal stress related to genocide and rape; 3) control - not directly exposed to genocide or rape. In contrast to previous work on similar samples, here we assess the influence of both prenatal exposures and postnatal experiences of adversity, such as adverse childhood experiences (ACEs), on first- and second-generation epigenetic age estimators. For our analysis, we hypothesized that adverse early life experiences would be associated with accelerated epigenetic age. For the prenatal adversity groups, we predict that single-exposed individuals will have a significantly accelerated epigenetic age compared to unexposed individuals, and double-exposed individuals will have a significantly accelerated epigenetic age relative to single-exposed and control individuals. We explored this relationship using first generation (Horvath, Hannum, PhenoAge) and second generation (GrimAge, DunedinPACE, YingDamAge, YingAdaptAge) epigenetic age estimators.

## RESULTS

Sample descriptive statistics are presented in Table 1. As noted earlier, all participants were 24 years of chronological age (mean= 24.1, sd = 0.10) and fairly evenly split by sex (n= 44 female, n=45 male). The majority (73.7%) had some college education. Reported adverse childhood events ranged from 1 to 11, with a mean of 5.00, and as previously reported, were highest in the doubly exposed group^2^.

**Table 1.** Descriptive Statistics.

|  | Double<br>Exposed<br>(N=30) | Single<br>Exposed<br>(N=31) | Control<br>(N=30) | Overall<br>(N=91) |
| --- | --- | --- | --- | --- |
| **Sex** |  |  |  |  |
| Female | 15 (50.0%) | 16 (51.6%) | 15 (50.0%) | 46 (50.5%) |
| Male | 15 (50.0%) | 15 (48.4%) | 15 (50.0%) | 45 (49.5%) |
| **Education Level** |  |  |  |  |
| primary level | 0 (0%) | 3 (9.7%) | 0 (0%) | 3 (3.3%) |
| secondary level | 29 (96.7%) | 20 (64.5%) | 25 (83.3%) | 74 (81.3%) |
| university level | 1 (3.3%) | 8 (25.8%) | 5 (16.7%) | 14 (15.4%) |
| **Socioeconomic Status** |  |  |  |  |
| Poorest | 6 (20.0%) | 2 (6.5%) | 0 (0%) | 8 (8.8%) |
| Poor | 17 (56.7%) | 14 (45.2%) | 4 (13.3%) | 35 (38.5%) |
| Middle Class | 7 (23.3%) | 15 (48.4%) | 26 (86.7%) | 48 (52.7%) |
| **ACEs Total** |  |  |  |  |
| Mean (SD) | 7.20 (2.12) | 5.74 (2.45) | 3.87 (2.29) | 5.60 (2.65) |
| Median [Min, Max] | 8.00 [4.00, 11.0] | 6.00 [1.00, 10.0] | 3.00 [1.00, 10.0] | 5.00 [1.00, 11.0] |

\*Single exposed - maternal stress related to genocide; \*\*Double-exposed - maternal stress related to genocide and rape; \*\*\*control - not directly exposed to genocide or rape;
Socioeconomic categories Income\*\*\*\* - Rwandans are classified into four socio-economic categories (Ibyiciro by'Ubudehe) by the Rwandan government based on income/consumption and household assets: Category 1: poorest, Category 2: poor, Category 3: middle class and Category 4: wealthy.

### Epigenetic age

Predicted epigenetic age ranges varied between clocks (see Supplemental Table 2). For example, the mean PhenoAge predicted age for the sample was 12.8 years (sd=5.61). We found no evidence of an association between epigenetic age acceleration and prenatal genocide exposure (single or double) and/or ACEs in the Hannum, Horvath, or PhenoAge clocks (Supplemental Tables 3.1- 3.6).

By contrast, epigenetic age acceleration calculated from YingDamAge, YingAdaptAge DundeinPACE, and GrimAgeAccel clocks were all associated with prenatal genocide exposure with stronger effects for the double-exposed group seen in the YingDamAge β*_single_* = 3.601, β*_doubl_*_e_= 6.375, *p* < 0.05) and YingAdaptAge clocks (β*_single_* = -6.482, β*_doubl_*_e_= -7.725, *p* < 0.001). The relationship between prenatal genocide exposure and epigenetic age acceleration calculated from YingAdaptAge remained significant and increased in both exposed groups (β*_single_* = -7.450, β*_double_* = -9.436, *p* < 0.001) even after controlling for ACEs. However, only the double-exposed group had significant epigenetic age acceleration calculated from YingDamAge after adjusting for ACEs, with the coefficient increasing after adjustment (β*_doubl_*_e_ =6.349, *p* < 0.01). The double-exposed group (but not single-exposed) was significantly associated with age acceleration in the DunedinPACE (β*_doubl_*_e_ = 0.048, *p* < 0.05) and GrimAgeAccel (β*_doubl_*_e_ = 1.448, *p* < 0.05) clocks. These effects were attenuated by adjustment for postnatal ACEs, especially in the DunedinPace and GrimAge2 clocks (see Tables 2.1 - 2.8). Visual examination of regression diagnostic plots in the performance package demonstrated good model performance for each second generation clock.

**Table 2.**
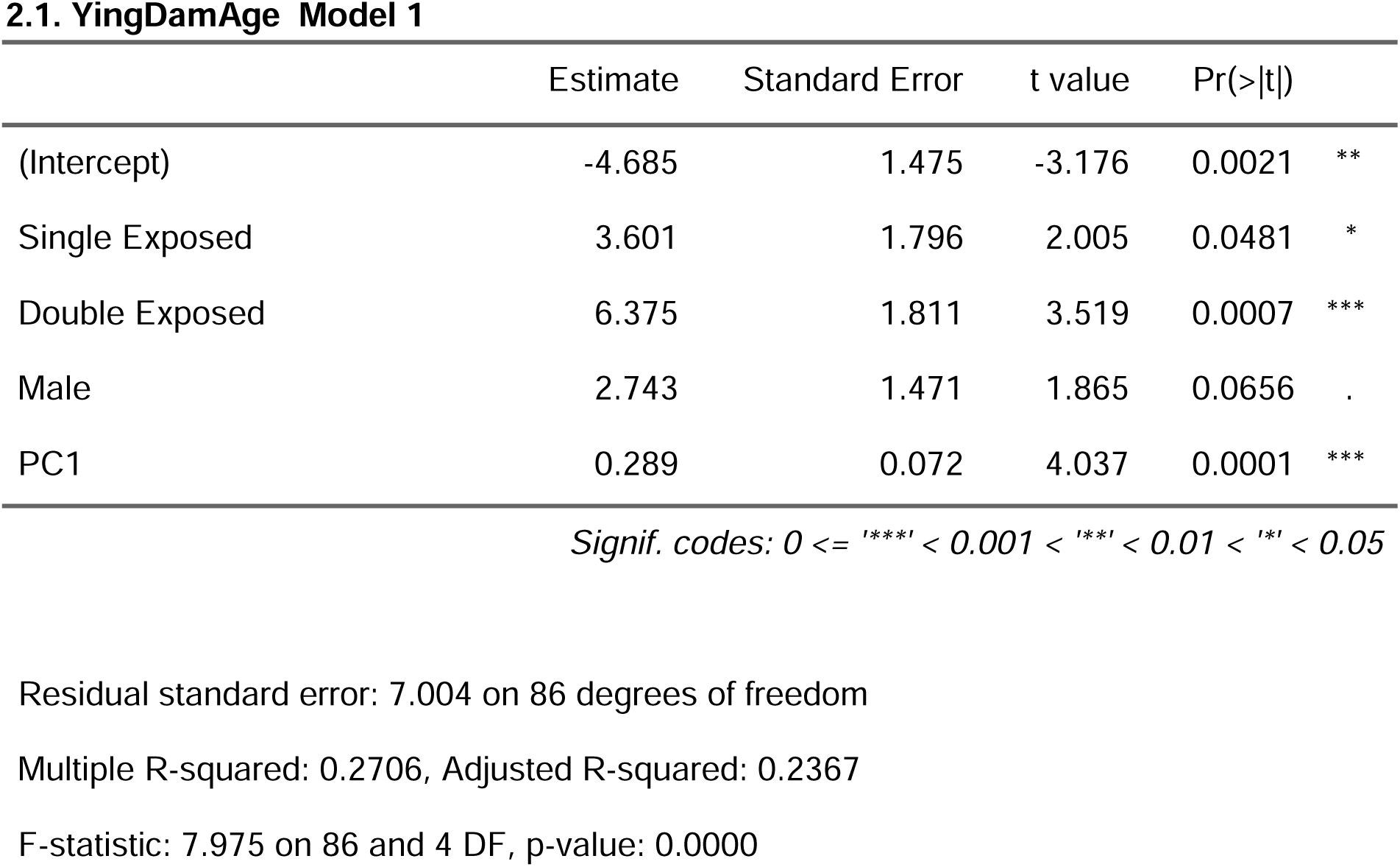
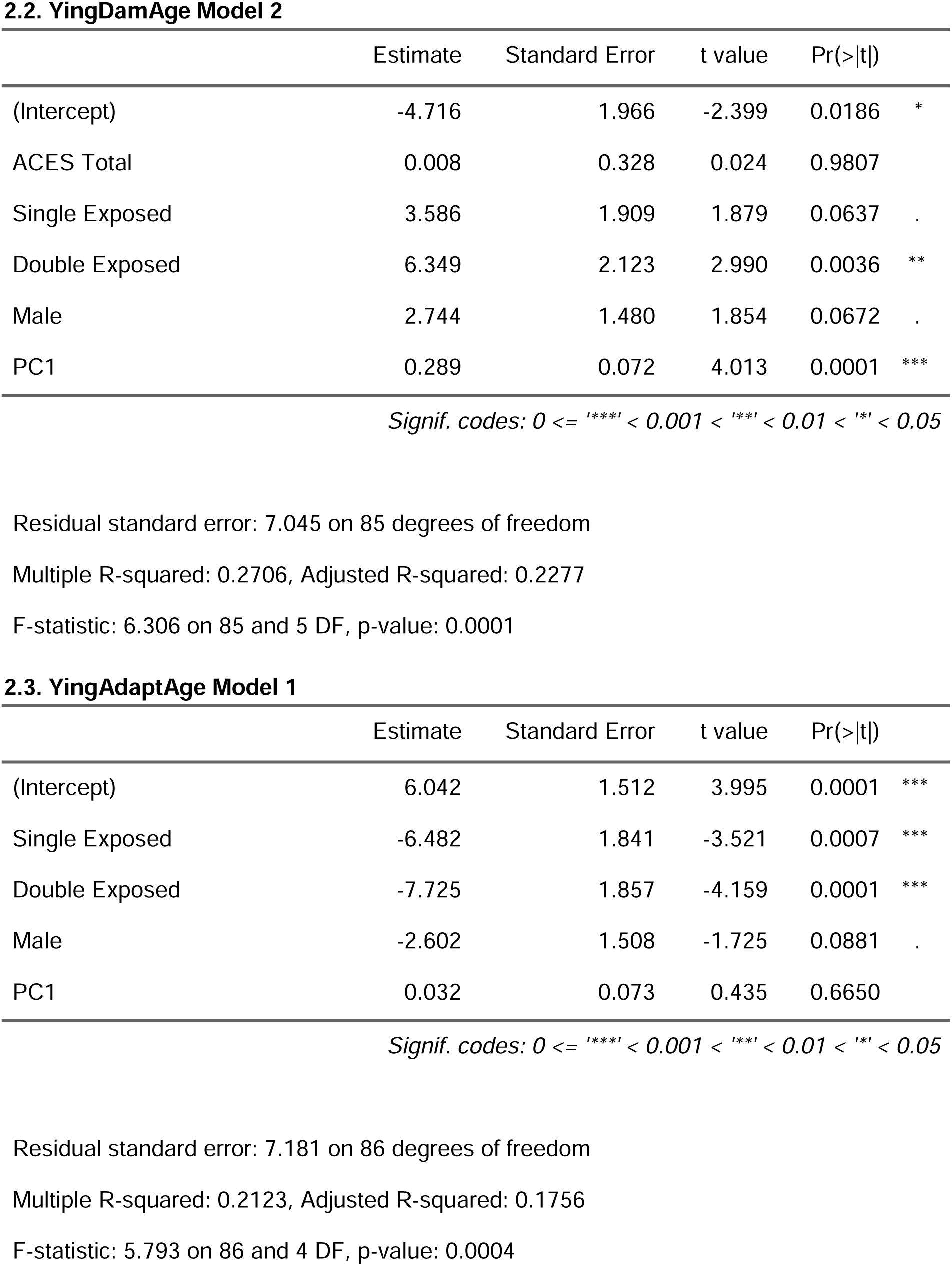
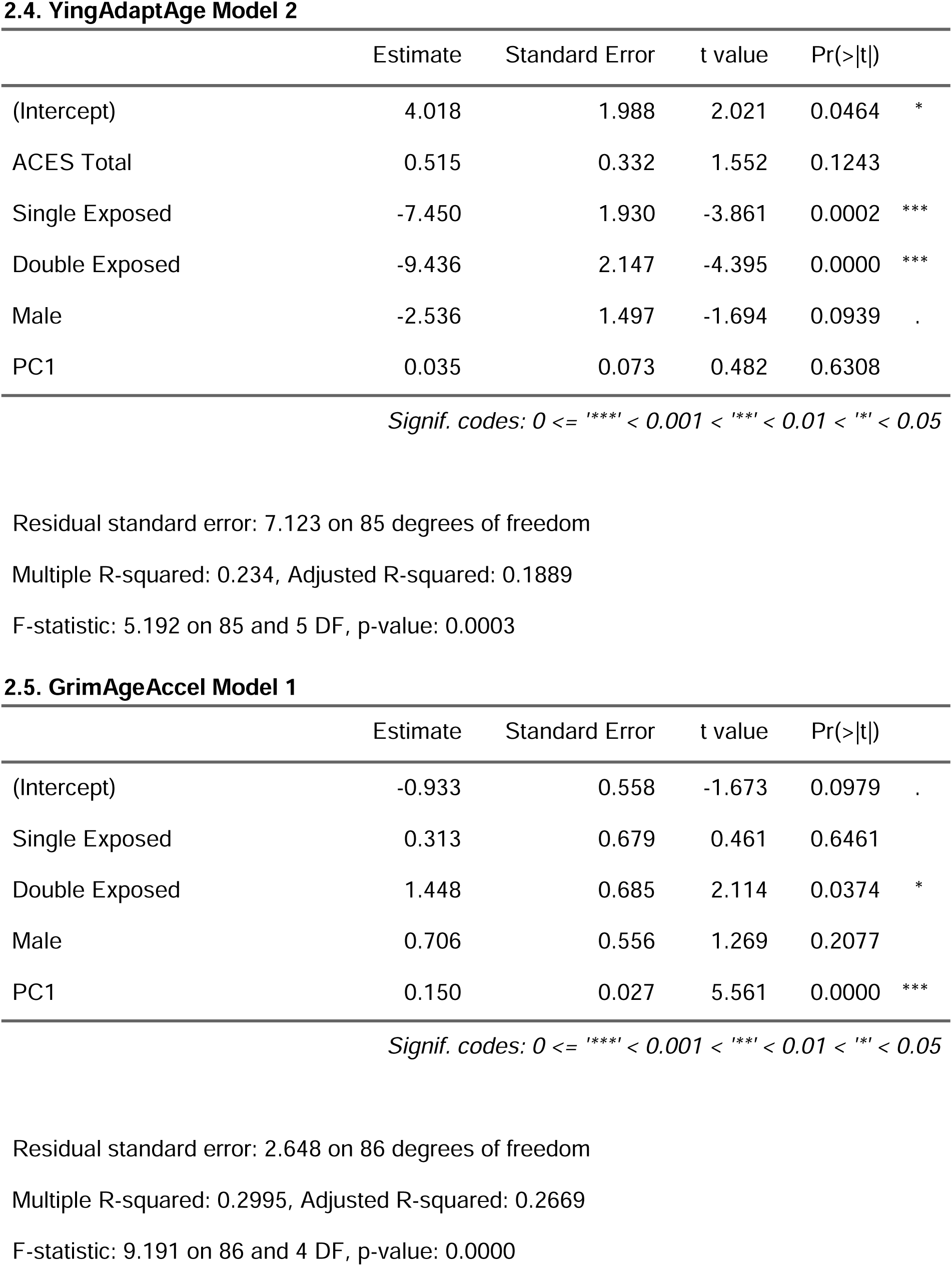
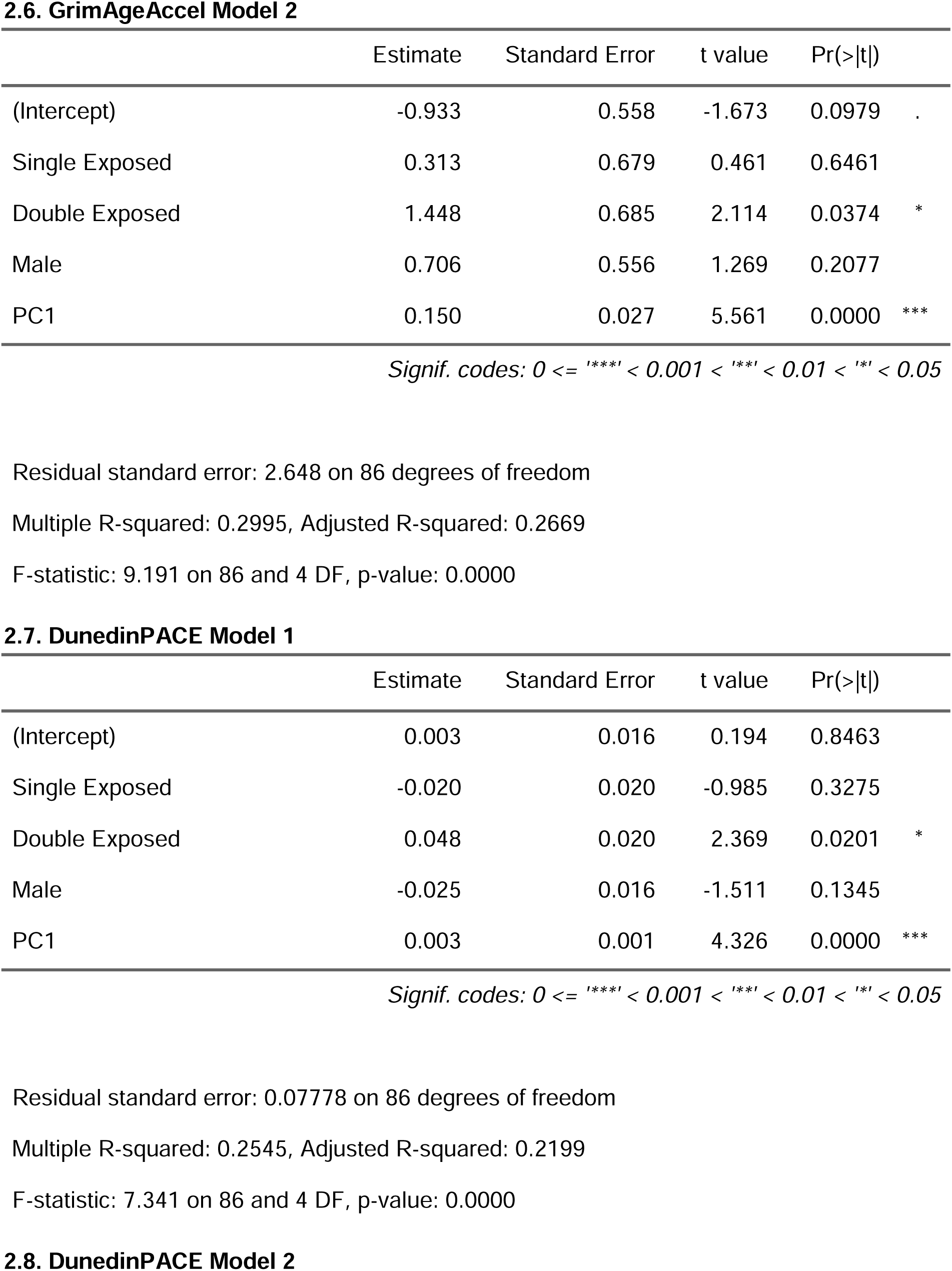
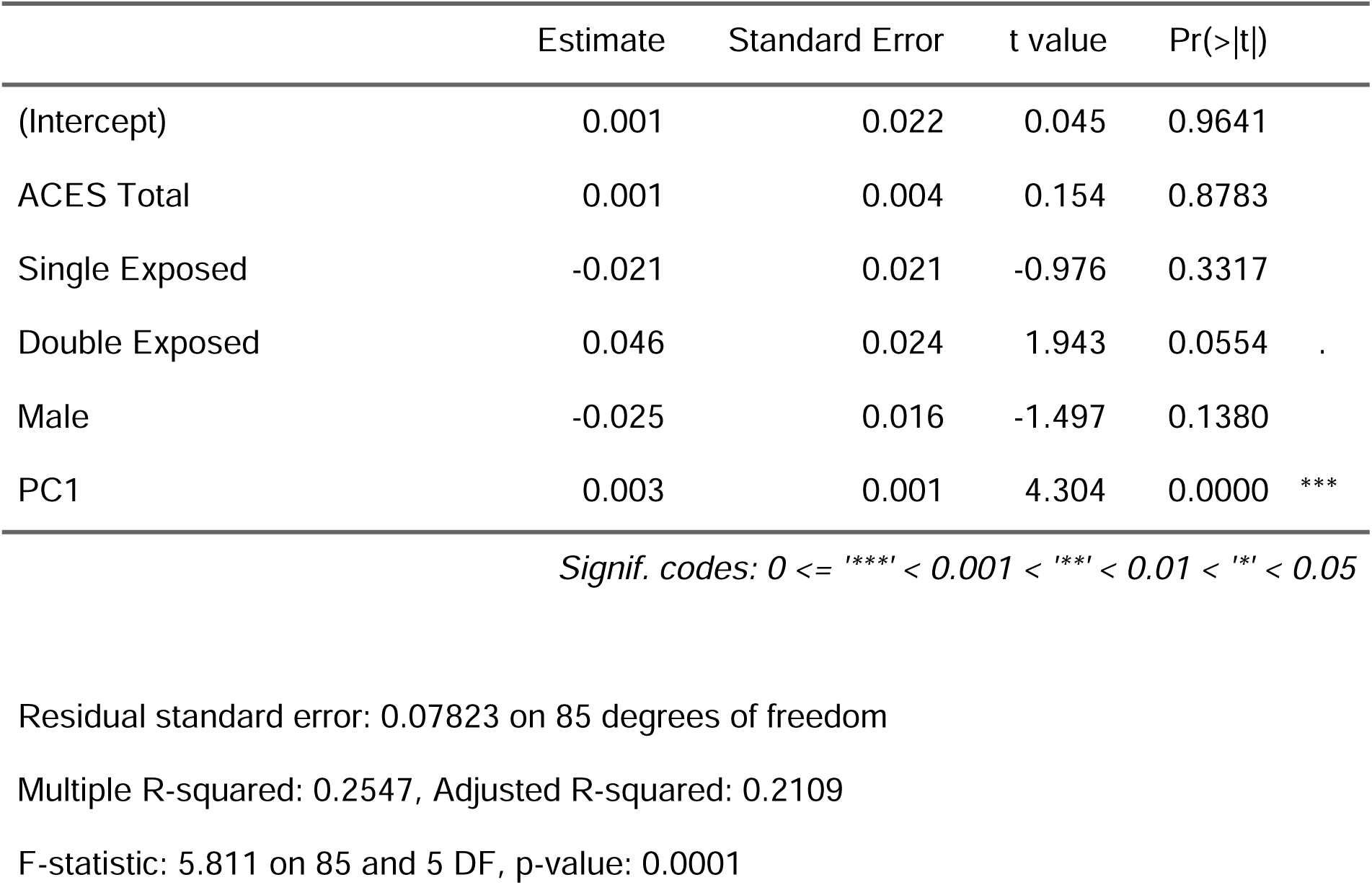
Second Generation Age Acceleration Models by Group.

We calculated the standardized mean difference to compare effect sizes across each of the models tested (Figure 1). The confidence intervals for each of the Horvath, Hannum, and PhenoAge models contained zero; in DunedinPACE and GrimAge, the single-exposed contrast, but not the double-exposed, also contained zero. Effect sizes were progressively greater (and greater for the double-exposed) in each second-generation clock tested. The largest effect size was for reduced adaptive aging in the double-exposed (YingAdaptAge SMD = -0.98, (95% CI = -1.54, -0.52) in Model 1 (unadjusted for ACEs) and Model 2 (YingAdaptAge SMD = - 1.19, 95% CI= -1.73, -0.66).

**Figure 1.** Forest plot comparing the effect of prenatal genocide exposure (vs. control) and epigenetic age acceleration across first and second generation epigenetic clocks. Comparison of effect sizes (standardized mean difference in age acceleration) across epigenetic age estimators and exposure groups. Model 1 is adjusted for sex and cell type; Model two is adjusted for sex, cell type, and ACEs.

## DISCUSSION

Previous research has demonstrated the sustained impacts of exposure to genocide and war-related trauma on the epigenome of individuals^4, 22, 24, 25^. Here, we reported the results of an analysis of epigenetic age among two groups of individuals conceived in Rwanda during the 1994 genocide and one group of individuals conceived outside of Rwanda during that time. We find a pattern of increased epigenetic aging between both prenatal exposure groups among second generation but not first generation epigenetic aging clocks, with stronger effects for the more severely exposed. Adjustment for postnatal adversity only slightly attenuated these effects, confirming the importance of prenatal developmental conditions. Our findings are provocative because they indicate that recent efforts to improve epigenetic clock construction by capturing physiological and/or causal variation in aging related DNA methylation may also better capture the impact of prenatal insults on healthspan. The effect sizes were greatest for the YingDamAge and YingAdaptAge, with the former increasing with adjustment for ACEs. While all of the second generation clocks were significantly different for the single-exposed group when adjusting for sex and immune cell composition, YingAdaptAge was the only measure that was significantly different once ACEs were added to the model. Similarly, effect sizes between exposure and epigenetic aging were greater in the second generation clocks, with the largest effect seen in reduced adaptive aging (YingAdaptAge) in the double-exposed. Links between prenatal conditions and earlier onset of age-related disease and decreased lifespan have been theorized within the Developmental Origins of Health and Disease (DOHaD) framework^26, 27^.

Low birthweight, prenatal exposure to stress and famine, and other indicators of poor developmental conditions have been associated with increased risk of chronic disease, decreased healthspan, and premature death in a well-established literature^28, 29^. Research exploring epigenetic age acceleration in young adults and associations with early life or prenatal conditions, however, is an emerging field; due to an individual’s limited ability to report on their own prenatal exposures, most studies in this field have investigated aging in children with parental report of exposure, or childhood adversity as self-reported by adults. Epigenetic age acceleration has been found in adults with in utero exposure to the Great Depression^30^, the Dutch Hunger Winter^31^, and other documented adverse developmental conditions. Our study adds to this literature by demonstrating the impact of prenatal exposure to genocide and genocidal rape in relatively healthy young adults. Whether these responses represent constraint, adaptation, or pathology is currently unclear, and should be a focus of study in future research^32^.

Our study also contributes to understanding some of the challenges and limitations of epigenetic clocks^33^, discerning causal relationships between epigenetic aging and healthspan, and the generalizability of epigenetic aging algorithms to non-Western populations^17^. We note, especially for the first generation clocks, the substantial variation in estimated age in these measures. The Hannum and PhenoAge clocks predicted age ranges between 2.40 - 32.1 years and 0.445 - 30.1 years, respectively. According to these predictions, some individuals had epigenetic age deceleration on the order of 23 years, which is remarkable considering the study participants were 24 years of chronological age at the time of assessment. It is also notable that outliers demonstrated epigenetic age *deceleration* and not acceleration and that the most extreme values were found in the double-exposed group. The Horvath clock had a more reasonable range (23.0, 38.3), with a slightly longer right tail to the distribution, reflecting more acceleration than deceleration results. It is worth noting that all the included epigenetic clocks are trained on reference datasets that are not representative of the Rwandan population in terms of ancestry and that were not exposed to such extreme prenatal and postnatal stress^17^.

As such, it is possible that these clocks are less accurate at estimating biological age in this population, potentially due to confounding by cell type^34–36^. For example, the Hannum clock was validated using a sample of 426 “Caucasian” and 230 “Hispanic” individuals, and the authors found a correlation of 96% between chronological and epigenetic age^15^. In our sample, this correlation was 3.9%. This finding highlights the limitations of creating epigenetic (or genetic, i.e. polygenic risk scores) algorithms based on non-fully representative datasets, reflecting broader anthropological critiques of biological normativity^37^.

First generation clocks, trained only on chronological age, may be particularly vulnerable to unmeasured confounding (e.g. population structure). The second generation clocks tested were also trained on data not representative of our sample (see Supplementary Table 1), but each include longitudinal measures of age-related decline, with the Ying clocks additionally constraining CpGs in its algorithm to those most causally related to age-related traits. Our findings suggest that these clocks are potentially more generalizable to other populations^17^.

### Limitations

We note several limitations to our study. The relatively small sample size may limit the generalizability of our findings. The majority of our participants had some college education which may not be representative of the socioeconomic status of our target population, but which we hypothesize would bias our results towards the null. Further, we utilized a cross-sectional study design with self-report on ACEs in adulthood, which introduces some degree of measurement error into our measure of ELA and limits causal inferences about the effect of ELA on biological aging. Finally, while we adjusted for ACEs, other unmeasured confounders, such as nutritional status, other environmental exposures, and genetic factors may have influenced the results. Despite these limitations, the shared directionality of effect between the multiple second-generation clocks tested, the highly impacted study population, and the use of an unexposed comparison group lend strength to our study findings.

## Conclusions

In sum, we found a relationship between maternal exposure to genocide-related trauma during pregnancy and epigenetic age acceleration in young adult offspring using second-generation but not first-generation epigenetic age estimators. Adjusting for adverse childhood experiences does not attenuate most of these results, and in the case of YingAdaptAge increased the coefficient. Future longitudinal research on larger samples should evaluate the potential interactive or moderating effects of both positive and negative environmental experiences later in the life course on epigenetic age acceleration.

## METHODS

This analysis is part of a comparative and associational cross-sectional study that explored the health impacts of prenatal exposure to genocide among Rwandan young adults conceived during the 100 days of genocide against the Tutsi in Rwanda in 1994 ^2, 10^. Study approvals were obtained from the Institutional Review Boards of the University of Illinois at Chicago (UIC: 2018- 1497), the University of Rwanda (UR No 063/CMHS IRB/2019), and Dartmouth College (STUDY0003231). All participants were given an information letter about the study and signed a ^2, 10^consent form before data collection. Rwandans aged 24 years old during the time of data collection were enrolled in the study and categorized into three groups accordion to their level of exposure: group 1: single-exposed - maternal stress-related genocide group 2: double-exposed - maternal stress-related genocide and rape; and group 3: control - not directly exposed to genocide or rape. The first participants in both exposed groups were recruited from the Solidarity for the Development of Widows and Orphans to Promote Self-Sufficiency and Livelihoods “SEVOTA” and Association of Genocide Widows Agahozo “AVEGA Agahozo”, non-profit organizations that support genocide survivors^2^. Participants in the group were descendants of Rwandans who were living outside the country during the time of the genocide and had no direct experience of the 1994 genocide. Each participant was invited to recommend age- and sex-matched Rwandans who belonged to any of the three groups.

Data were collected by the first author, who is a Rwandan mental health nurse, in a private room. Interviews were conducted in Kinyarwanda. A total of 91 participants completed demographic and health-related surveys in Research Electronic Data Capture (REDCap).

### Prenatal exposure to genocide

To determine the level of exposure, we asked each participant if they were conceived by a genocide survivor and whether they were conceived via genocidal rape. Most of the participants in the exposed groups were referred to the study by an organization that supports survivors of genocide and their offspring; these organizations hence knew and shared with us in which category their referred potential participants belonged to. We verified these exposures with participants during their interviews. We conducted screening interviews with participants in the control group to determine if individuals were born to Tutsi women who were living outside the country during the time of the genocide. We excluded participants if their parents left the country due to the genocide or other political unrest in the months leading up to the genocide. We backdated participants’ dates of birth to estimate if they were conceived during the time of the genocide: April 07 - July 4, 1994. An equal number of female and male participants were enrolled in each group (Table 1).

### Early Life Adversity

Adverse childhood experiences before age 18 were assessed using the Adverse Childhood Experiences International Questionnaire (ACEs IQ)^38^. This measure includes 13 items - emotional abuse; physical abuse; sexual abuse; violence against household members; living with household members who were substance abusers; living with household members who were mentally ill or suicidal; living with household members who were imprisoned; one or no parents, parental separation or divorce; emotional neglect; physical neglect; bullying; community violence; and collective violence, resulting in an ACEs score of 0-13. This measure has been validated in another African setting^39^ and had acceptable internal consistency within our sample (α = 0.70).

### Dried Blood Spot Collection

Dried blood spots (DBS) were collected for later DNA methylation analysis following the interview. DBS is a minimally invasive and frequently used method that is convenient and cost- effective in research in remote settings that requires long-distance transportation of samples^40^. The use of DBS has been validated in studies exploring DNAm ^41^ (24). Whole blood drops were collected from a finger stick by sterile lancet on Flinders Technology Associates cards (FTA), with four sample areas 125 μL each per card. Samples were collected from March 07 to April 06, 2019. The drops were air-dried for at least four hours before placing each card in an airtight envelope with silica-based desiccant and stored at room temperature in Rwanda. Samples were then shipped to the University of Illinois on April 7, 2019, and later to Dartmouth College on August 26, 2021, where they were stored at -80 °C prior to sample processing and DNA methylation analysis in April 2022.

### DNA Methylation Sample Processing

DNA was extracted from dried blood spots (DBS) using QIAamp DNA Investigator Kit (Qiagen, Catalog #56504). The manufacturer’s protocol was optimized to improve DNA yield. For each sample (N = 91), two 6 mm hole punches were processed in individual 1.5 microcentrifuge tubes (Eppendorf) and QIAamp MinElute columns (Qiagen). The elution buffer ATE (Qiagen) was heated to 70°C to improve the release of DNA from the silica membrane. 60 µL of ATE were pipetted onto the silica membrane of the MinElute column and incubated at room temperature (15–25°C) for 10 min. before centrifugation. The eluate was re-eluted onto the silica membrane and incubated at room temperature (15–25°C) for 3 min. Following final centrifugation, the eluates were combined into one 1.5 microcentrifuge tube and carefully pipetted up and down to ensure sufficient mixture. Purified DNA was quantified using Invitrogen Qubit 3.0 Fluorometer broad range assay (median = 259.6 ng of DNA). Infinium FFPE QC and DNA Restoration kit (Illumina Inc., WG-321-1001, WG-321-1002) was used to evaluate sample quality and restore degraded DNA, prior to bisulfite treatment (Zymo EZDNA Methylation Kit, Zymo Research, Irvine, CA, USA). DNA methylation was measured using MethylationEPIC v.1 beadchip, and samples were randomized by the prenatal exposure group across chips.

### Data Preprocessing and Normalization

DNA methylation microarray idat files were imported into R (version 4.2.1) and processed using the *minfi* package (version 1.41.0)^42^. Quality control included estimating sex and calculating mean detection p-value for CpGs across all samples to evaluate signal reliability. Beta and M values were calculated using the normal-exponential out-of-band (Noob) method, recommended for the 12-immune-cell-type extended deconvolution, which includes normalization and background correction^36^. Further preprocessing took place before any epigenomic analysis, including filtering CpGs with low detection p-value across samples (20,170), filtering probes on X and Y chromosomes (Y= 135, X = 18,588), and filtering SNPs/cross-hybridizing probes β-values and M-values were extracted and used in subsequent analysis.

### Epigenetic age acceleration

All participants were 24 years of age during data collection. Epigenetic age was calculated from the preprocessed methylation beta values across all samples using the methyAge function in package ENmix (version 1.32.0), which includes *Horvath, Hannum,* and *PhenoAge* clocks.

*DunedinPACE, GrimAgeAccel*, *YingAdaptage,* and *YingDamAge* were similarly calculated using the *Biolearn* library developed and maintained by the Biomarkers of Aging Consortium^43^. While several of these clocks were developed using the Illumina 450K array, each has been validated for use with the EPIC microarray used in the present analysis^44^. After calculating the predicted epigenetic age, we regressed chronological age on the predicted age (except for GrimAgeAccel, which produces its own residualized score), and the age-corrected residual difference was used in all analyses as our epigenetic age acceleration measure.

### Immune cell type deconvolution

Immune cell type proportions were estimated across all samples using a DNA methylation-derived cell type deconvolution method. The package *FlowSorted.BloodExtended.EPIC* (version 2.0.0) infers proportions of 12 immune cell types; neutrophils (Neu), monocytes (Mono), basophils (Bas), eosinophils (Eos), CD4T naïve cells (CD4nv), CD4T memory cells (CD4mem), B naïve cells (Bnv), B memory cells (Bmem), CD8T naïve cells (CD8nv), CD8T memory cells (CD8mem), T regulatory cells (Treg), and natural killer cells (NK)^36^. The monocyte-to-lymphocyte and neutrophil-to-lymphocyte ratios were calculated based on the deconvoluted cell proportions and were used as primary outcomes. To summarize immunophenotype, a principal component analysis was conducted on all cell types; the first principal component explained 68.2% of the variance and was used to control for cell-type composition. The first immune cell type principal component (PC) was included as a covariate in the epigenetic age analysis.

### Data Analysis

All statistical analyses were conducted in R version 4.3.1. Descriptive statistics were run across the exposure groups before analysis, and epigenetic aging measures were plotted by the exposure group and visually inspected (see Supp Fig 1a & 1b). All participants had complete data. For each epigenetic clock, we conducted a two-step multiple linear regression to determine the relationship between the exposure group and epigenetic age acceleration, with the control group serving as the reference category. In the first step, we adjusted for sex and the first immune cell-type PC; in the second, we added the ACE total score as an additional predictor alongside the exposure group. We also produced overlapping density plots and within-individual Pearson correlations between the clocks to assess the consistency of the predictions (Supplemental Figure 2 & 3). To compare effects across different epigenetic clocks, we calculated the standard mean difference of the regression coefficients for single or double- exposed group status vs. control contrast for models and plotted them together in a forest plot (see Figure 1).

## Data Availability

Data are available for replication purposes but not for new research as per community requests and the informed consent process.

## Additional Information

The authors declare no competing interests.

