## Supplemental Files for "Prenatal exposure to genocide accelerates epigenetic aging as measured in second-generation clocks among young adults"

**Supplemental Information**

Supplemental Table 1. Overview of Epigenetic Clock Construction	1

Supplemental Table 2. Predicted Epigenetic Ages	3

Supplemental Figure 1a. Age Acceleration by Exposure Group	5

Supplemental Figure 1b. Page of aging by exposure group	6

Supplemental Figure 2. Overlapping Predicted Age Density Plot	7

Supplemental Figure 3.a Intra Clock Correlation Plot	8

Supplemental Figure 3b. Correlations between Predicted and Chronological Age	9

**Supplementary Table 4: First Generation Age Acceleration Models by Group**	10

S4.1. Horvath Model 1	10

S4.2. Horvath Model 2	10

S4.3. Hannum Model 1	11

S4.4. Hannum Model 2	11

S4.5. PhenoAge Model 1	11

S4.6. PhenoAge Model 2	12

##### Supplemental Table 1. Overview of Epigenetic Clock Construction

| Name | Training data | Additional Phenotypes | #CpG |
| --- | --- | --- | --- |
| Horvath | 7,844 samples from non cancer datasets, largely from USA/Europe  Age range 0 -101  Pan tissue  Illumina 450k  Illumina 27K | none | 353 |
| Hannum | 426 Caucasian and 230 Hispanic individuals,  Age range 19 - 101  Venous blood  Illumina 450K | - Gender - BMI | 71 |
| PhenoAge | ~16k NHANES participants provided biomarker and chronological age data to train Model 1.  Model 1 + DNAm data in 456 individuals living in Tuscany (Invecchiare study) used to create PhenoAge.  Age range 21 - 100  Venous blood  Illumina 450K  Illumina 27K | - Albumin - Creatinine - Glucose - CRP - Lymphocyte percent - Mean cell volume - Red cell distribution width - alkaline phosphatase - white blood cell count | 513 |
| GrimAgeAccel | 2,356 individuals from the Framingham heart study (FHS) Offspring Cohort  Mean age 66.5  Venous blood  Illumina 450K | - DNAm pack-years - Age - Sex - DNAm-based surrogate markers of plasma proteins:   - adrenomedullin (ADM)   - beta-2-microglobulim (B2M)   - cystatin C (Cystatin C)   - GDF-15, leptin (Leptin)   - PAI-1   - tissue inhibitor metalloproteinases 1 (TIMP-1) - Time to death (all cause mortality) | 1030 |
| DunedinPACE | 1037 predominantly white individuals from The Dunedin Study (New Zealand)  Age range 26 - 45  Venous blood  Illumina 450K | Decline between two timepoints approximately 10 years apart in:   - albumin - alkaline phosphatase - blood urea nitrogen - creatinine - C-reactive protein - HbA1C - systolic blood pressure - forced expiratory volume in one second | 173 |
| YingDamAge & AdaptYingAge | 27,750 European individuals from 36 different cohorts.  DamAge CpGs are associated with poorer aging  AdaptAge CpGs are associated with longevity and healthy aging | Epigenome-wide Mendelian randomization identified CpGs most causally associated with 12 age related traits:   - lifespan - extreme longevity - healthspan - frailty index - self-rated health - Horvath age - Hannum age - PhenoAge - GrimAge - Aging-GIP1 (e.g. estimated genetic contribution to aging) - socioeconomic traits-adjusted Aging-GIP1 - healthy aging |  |

##### Supplemental Table 2. Predicted Epigenetic Ages

|  | Double Exposed (N=30) | Single Exposed (N=31) | Control (N=30) | Overall (N=91) |
| --- | --- | --- | --- | --- |
| **Chronological Age** |  |  |  |  |
| Mean (SD) | 24.1 (0.104) | 24.2 (0.0963) | 24.1 (0.0814) | 24.1 (0.0957) |
| Median [Min, Max] | 24.1 [24.0, 24.5] | 24.2 [24.0, 24.3] | 24.1 [24.0, 24.3] | 24.1 [24.0, 24.5] |
| **Horvath** |  |  |  |  |
| Mean (SD) | 30.0 (3.47) | 30.2 (3.48) | 30.0 (3.56) | 30.1 (3.46) |
| Median [Min, Max] | 30.2 [23.8, 36.7] | 30.1 [23.8, 38.3] | 30.0 [23.0, 37.8] | 30.1 [23.0, 38.3] |
| **Hannum** |  |  |  |  |
| Mean (SD) | 23.8 (6.05) | 24.9 (4.08) | 25.5 (3.08) | 24.7 (4.57) |
| Median [Min, Max] | 24.7 [2.40, 32.1] | 25.3 [15.0, 31.9] | 26.3 [19.1, 30.5] | 25.2 [2.40, 32.1] |
| **PhenoAge** |  |  |  |  |
| Mean (SD) | 13.5 (5.75) | 12.3 (6.21) | 12.8 (4.77) | 12.9 (5.57) |
| Median [Min, Max] | 14.0 [0.445, 30.1] | 13.5 [0.576, 23.0] | 13.8 [1.14, 18.7] | 13.8 [0.445, 30.1] |
| **DunedinPACE** |  |  |  |  |
| Mean (SD) | 1.04 (0.0948) | 0.989 (0.0715) | 0.999 (0.0932) | 1.01 (0.0889) |
| Median [Min, Max] | 1.02 [0.849, 1.30] | 0.992 [0.833, 1.12] | 0.983 [0.773, 1.22] | 1.00 [0.773, 1.30] |
| **GrimAgeAccel** |  |  |  |  |
| Mean (SD) | 0.663 (3.47) | -0.0965 (2.96) | -0.564 (2.79) | 0.0000000000220 (3.09) |
| Median [Min, Max] | 0.0184 [-4.94, 13.1] | 0.196 [-6.63, 5.66] | -0.847 [-6.16, 7.18] | 0.0390 [-6.63, 13.1] |
| **YingDamAge** |  |  |  |  |
| Mean (SD) | 34.7 (8.34) | 33.5 (8.09) | 28.9 (7.13) | 32.4 (8.17) |
| Median [Min, Max] | 34.1 [17.5, 50.1] | 33.9 [20.8, 48.9] | 27.7 [15.9, 42.5] | 31.4 [15.9, 50.1] |
| **YingAdaptAge** |  |  |  |  |
| Mean (SD) | 17.6 (7.89) | 18.3 (5.83) | 25.2 (8.02) | 20.3 (8.00) |
| Median [Min, Max] | 19.3 [-3.35, 29.4] | 18.8 [5.94, 27.6] | 25.2 [9.06, 39.9] | 20.2 [-3.35, 39.9] |

##### Supplemental Figure 1a. Age Acceleration by Exposure Group

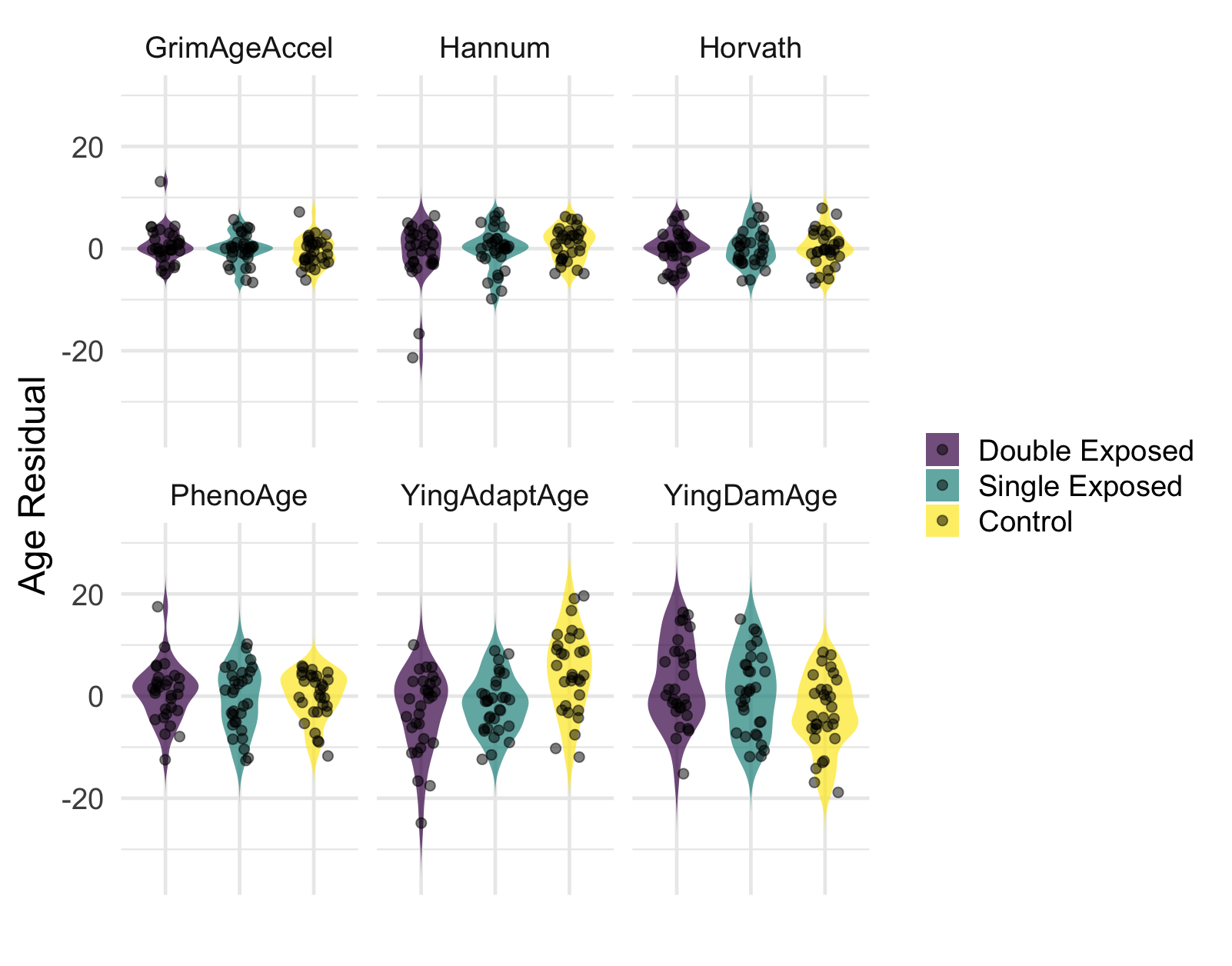

##### Supplemental Figure 1b. Page of aging by exposure group

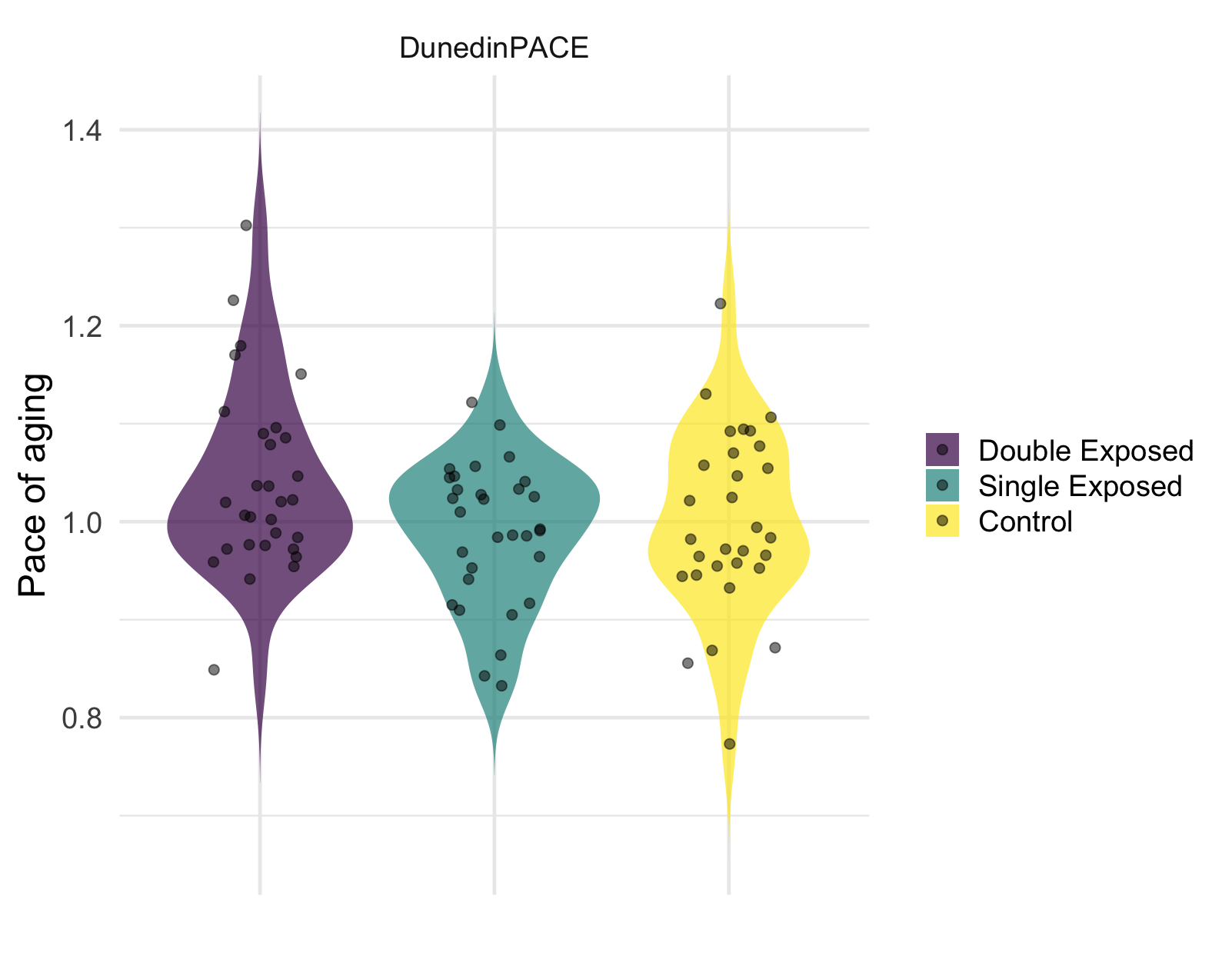

##### Supplemental Figure 2. Overlapping Predicted Age Density Plot

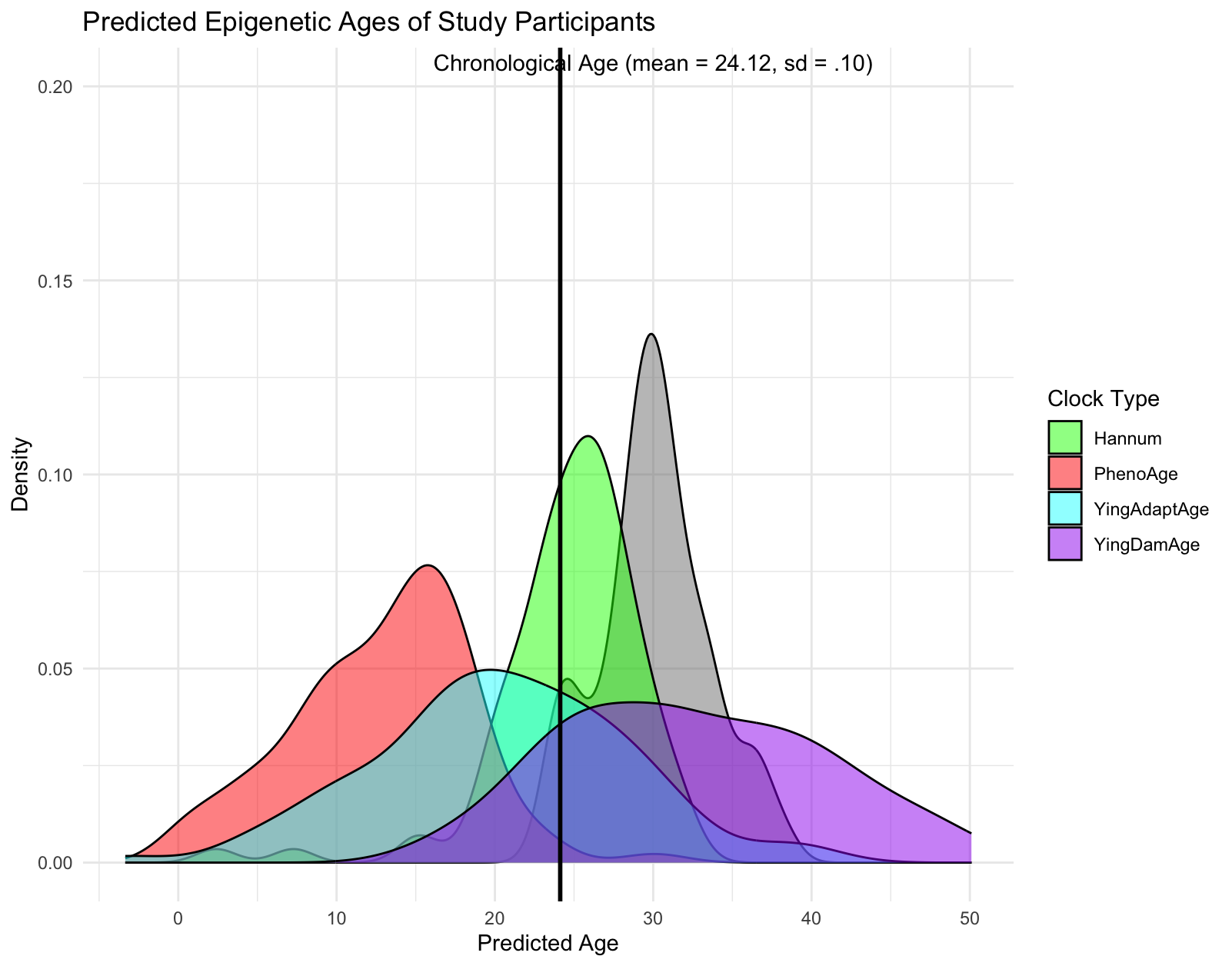

##### Supplemental Figure 3.a Intra Clock Correlation Plot

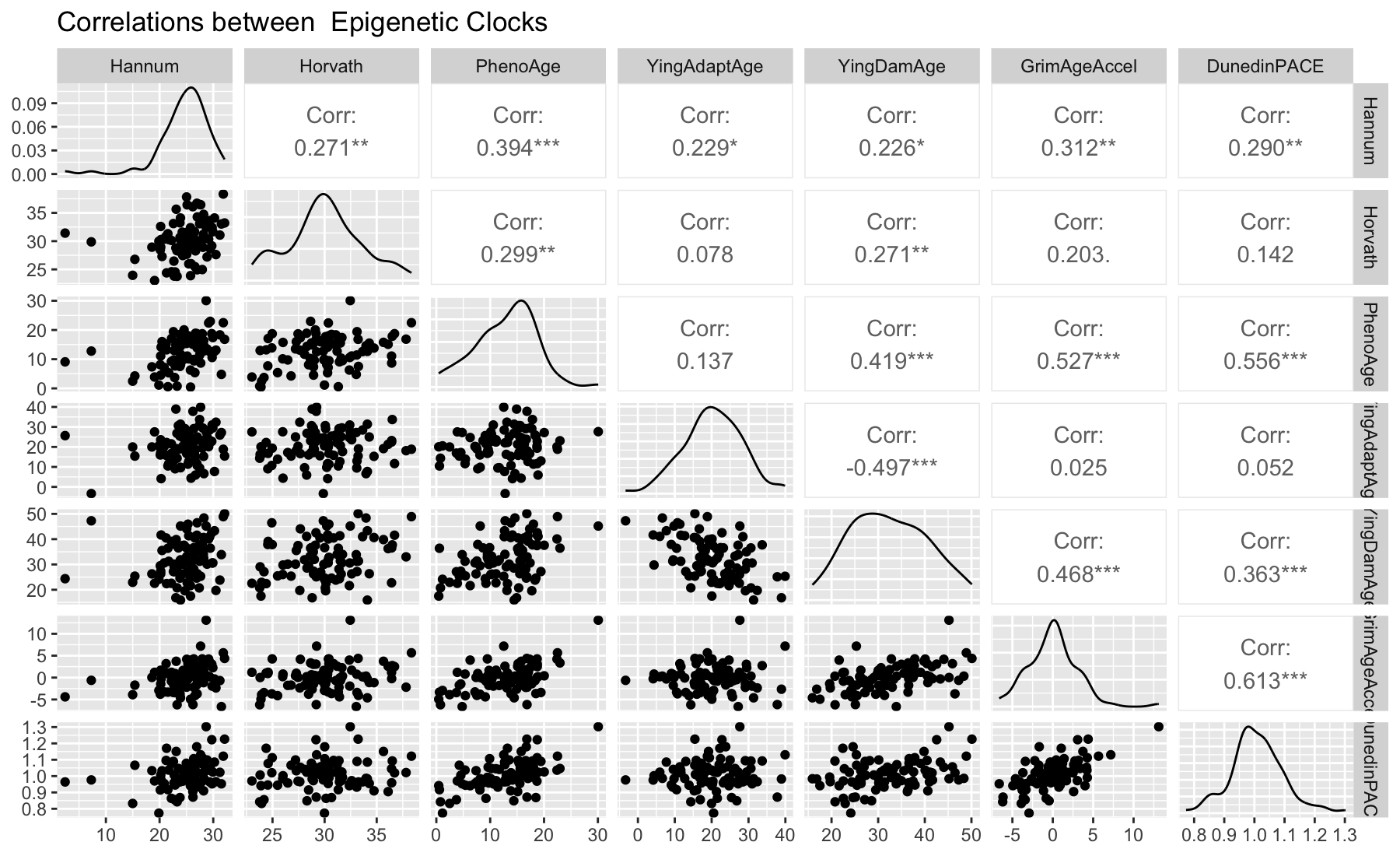

##### Supplemental Figure 3b. Correlations between Predicted and Chronological Age

(DunedinPACE and GrimAgeAccel excluded as only pace of aging and age acceleration is estimated)

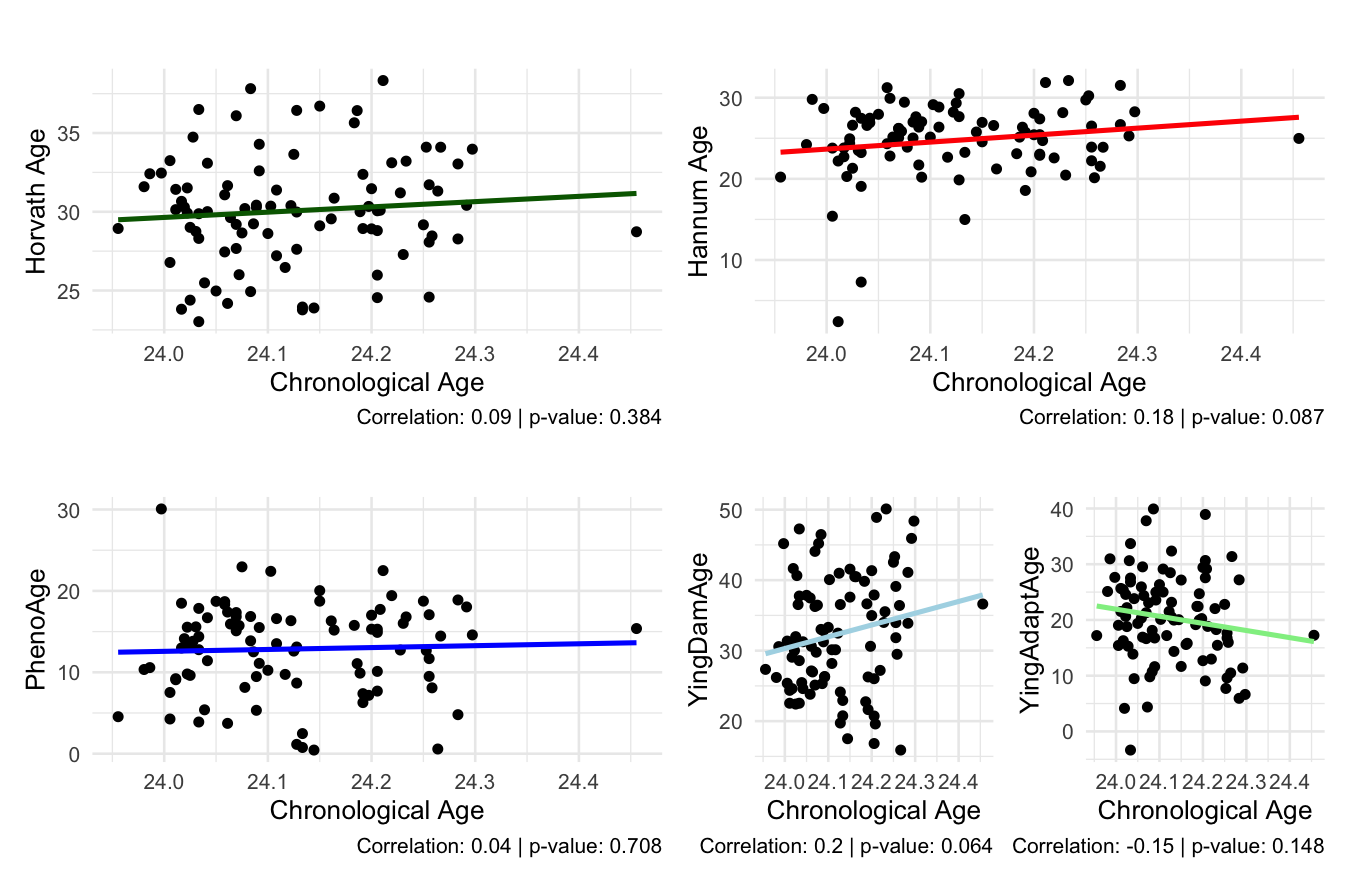

##### Supplementary Table 3: First Generation Age Acceleration Models by Group

##### **S3.1.** **Horvath Model 1**

|  | Estimate | Standard Error | t value | Pr(>\|t\|) |
| --- | --- | --- | --- | --- |
| (Intercept) | -0.565 | 0.734 | -0.770 | 0.4435 |
| Single Exposed | 0.165 | 0.894 | 0.184 | 0.8542 |
| Double Exposed | -0.003 | 0.902 | -0.004 | 0.9970 |
| Male | 1.032 | 0.732 | 1.409 | 0.1624 |
| PC1 | -0.013 | 0.036 | -0.367 | 0.7143 |
| *Signif. codes: 0 <= '***' < 0.001 < '**' < 0.01 < '*' < 0.05* | | | | |
| Residual standard error: 3.487 on 86 degrees of freedom | | | | |
| Multiple R-squared: 0.02376, Adjusted R-squared: -0.02164 | | | | |
| F-statistic: 0.5233 on 86 and 4 DF, p-value: 0.7188 | | | | |

##### **S3.2.** **Horvath Model 2**

|  | Estimate | Standard Error | t value | Pr(>\|t\|) |
| --- | --- | --- | --- | --- |
| (Intercept) | -0.916 | 0.977 | -0.937 | 0.3513 |
| ACEs Total | 0.089 | 0.163 | 0.547 | 0.5861 |
| Single Exposed | -0.003 | 0.949 | -0.003 | 0.9977 |
| Double Exposed | -0.300 | 1.055 | -0.284 | 0.7771 |
| Male | 1.044 | 0.736 | 1.418 | 0.1597 |
| PC1 | -0.013 | 0.036 | -0.350 | 0.7272 |
| *Signif. codes: 0 <= '***' < 0.001 < '**' < 0.01 < '*' < 0.05* | | | | |
| Residual standard error: 3.501 on 85 degrees of freedom | | | | |
| Multiple R-squared: 0.02718, Adjusted R-squared: -0.03004 | | | | |
| F-statistic: 0.475 on 85 and 5 DF, p-value: 0.7939 | | | | |

##### **S3.3.** **Hannum Model 1**

|  | Estimate | Standard Error | t value | Pr(>\|t\|) |  |
| --- | --- | --- | --- | --- | --- |
| (Intercept) | 0.774 | 0.936 | 0.827 | 0.4103 |  |
| Single Exposed | -1.056 | 1.140 | -0.926 | 0.3568 |  |
| Double Exposed | -1.500 | 1.150 | -1.305 | 0.1954 |  |
| Male | 0.161 | 0.934 | 0.173 | 0.8633 |  |
| PC1 | 0.090 | 0.045 | 1.992 | 0.0496 | * |
| *Signif. codes: 0 <= '***' < 0.001 < '**' < 0.01 < '*' < 0.05* | | | | | |
| Residual standard error: 4.445 on 86 degrees of freedom | | | | | |
| Multiple R-squared: 0.06622, Adjusted R-squared: 0.02278 | | | | | |
| F-statistic: 1.525 on 86 and 4 DF, p-value: 0.2022 | | | | | |

##### **S3.4.** **Hannum Model 2**

|  | Estimate | Standard Error | t value | Pr(>\|t\|) |  |
| --- | --- | --- | --- | --- | --- |
| (Intercept) | 0.560 | 1.247 | 0.449 | 0.6548 |  |
| ACEs Total | 0.055 | 0.208 | 0.263 | 0.7934 |  |
| Single Exposed | -1.159 | 1.211 | -0.957 | 0.3414 |  |
| Double Exposed | -1.682 | 1.347 | -1.249 | 0.2152 |  |
| Male | 0.168 | 0.939 | 0.179 | 0.8581 |  |
| PC1 | 0.091 | 0.046 | 1.988 | 0.0501 | . |
| *Signif. codes: 0 <= '***' < 0.001 < '**' < 0.01 < '*' < 0.05* | | | | | |
| Residual standard error: 4.469 on 85 degrees of freedom | | | | | |
| Multiple R-squared: 0.06697, Adjusted R-squared: 0.01209 | | | | | |
| F-statistic: 1.22 on 85 and 5 DF, p-value: 0.3067 | | | | | |

##### **S3.5.** **PhenoAge Model 1**

|  | Estimate | Standard Error | t value | Pr(>\|t\|) |  |
| --- | --- | --- | --- | --- | --- |
| (Intercept) | -0.086 | 1.063 | -0.081 | 0.9360 |  |
| Single Exposed | -0.856 | 1.294 | -0.661 | 0.5103 |  |
| Double Exposed | 1.045 | 1.306 | 0.801 | 0.4255 |  |
| Male | 0.066 | 1.060 | 0.062 | 0.9508 |  |
| PC1 | 0.244 | 0.052 | 4.733 | 0.0000 | *** |
| *Signif. codes: 0 <= '***' < 0.001 < '**' < 0.01 < '*' < 0.05* | | | | | |
| Residual standard error: 5.048 on 86 degrees of freedom | | | | | |
| Multiple R-squared: 0.2146, Adjusted R-squared: 0.178 | | | | | |
| F-statistic: 5.873 on 86 and 4 DF, p-value: 0.0003 | | | | | |

##### **S3.6.** **PhenoAge Model 2**

|  | Estimate | Standard Error | t value | Pr(>\|t\|) |  |
| --- | --- | --- | --- | --- | --- |
| (Intercept) | -0.495 | 1.415 | -0.350 | 0.7274 |  |
| ACEs Total | 0.104 | 0.236 | 0.441 | 0.6604 |  |
| Single Exposed | -1.051 | 1.374 | -0.765 | 0.4463 |  |
| Double Exposed | 0.699 | 1.529 | 0.458 | 0.6484 |  |
| Male | 0.079 | 1.066 | 0.074 | 0.9410 |  |
| PC1 | 0.245 | 0.052 | 4.722 | 0.0000 | *** |
| *Signif. codes: 0 <= '***' < 0.001 < '**' < 0.01 < '*' < 0.05* | | | | | |
| Residual standard error: 5.072 on 85 degrees of freedom | | | | | |
| Multiple R-squared: 0.2163, Adjusted R-squared: 0.1703 | | | | | |
| F-statistic: 4.693 on 85 and 5 DF, p-value: 0.0008 | | | | | |
